# Neighborhood Socioeconomic Status and Healthcare Quality Measures Following Bariatric Surgery in Maryland

**DOI:** 10.1101/2025.09.18.25336081

**Authors:** Oluwasegun Akinyemi, Terrence Fullum, Mojisola Fasokun, Nkemdirim Ugochukwu, Kakra Hughes, Dahai Yue, Craig Scott Fryer, Jie Chen, Kellee White-Whilby

**Author notes:** Corresponding author: Oluwasegun Akinyemi.

## Abstract

**Introduction:** Bariatric surgery is the most effective treatment for severe obesity, yet significant socioeconomic disparities in access and outcomes persist, especially in disadvantaged communities. Neighborhood socioeconomic status (nSES) influences healthcare utilization, complication rates, and recovery, but its impact within state-specific reimbursement models remains understudied.

**Objective:** This study examines whether nSES, measured by the Distressed Communities Index (DCI), is independently associated with prolonged hospital stays and higher readmission rates after bariatric surgery. It also evaluates the interaction between race/ethnicity and nSES, to assess disparities in post-surgical outcomes.

**Methodology:** A retrospective cohort study was conducted using Maryland State Inpatient Databases (SID) from 2018 to 2020. The study population included all adult patients who underwent bariatric surgery, identified using ICD-10 procedure codes. The primary explanatory variable was nSES, operationalized using DCI quintiles, linked to patient ZIP codes. The primary outcomes were hospital length of stay (continuous variable) and readmission (binary variable: Yes/No). Multivariate linear regression (for hospital stay) and logistic regression (for readmission) were performed, adjusting for demographic factors (age, sex, race/ethnicity), clinical characteristics (preexisting comorbidities using the Charlson Comorbidity Index (CCI) and, obesity class), and socioeconomic indicators (insurance type) and Geographic classification. Interaction terms were included to evaluate whether race/ethnicity modified the association between nSES and these outcomes.

**Result:** Among 10,784 bariatric surgery recipients, the majority were Black (48.3%), female (83.1%), with a mean age of 44.1 ± 11.6 years. Length of stay did not differ significantly by DCI Quintiles; patients in distressed areas had similar odds of prolonged hospitalization compared to those in prosperous areas (β = 0.045; 95% CI: –0.111 to 0.201; p = 0.575). Readmission risk was higher in distressed neighborhoods (OR = 1.64; 95% CI: 0.76–3.54; p = 0.207), though not statistically significant. No interaction was observed between nSES and race/ethnicity.

**Conclusion:** Residents of disadvantaged neighborhoods showed a non-significant trend toward higher readmission without increased hospital stay. Findings underscore the need to enhance post-discharge care for socioeconomically vulnerable populations.

## INTRODUCTION

Bariatric surgery is the most effective long-term intervention for severe obesity, leading to sustained weight loss and improvement in obesity-related comorbidities such as diabetes and hypertension(1–3). However, post-operative healthcare quality measures, including hospital length of stay (LOS) and readmission rates, serve as critical indicators of patient outcomes and healthcare system efficiency and post-surgical recovery(4, 5). Prolonged hospital stays increase healthcare costs and indicate potential complications, while readmissions often reflect suboptimal perioperative care, surgical complications, or social barriers to recovery(6, 7).

Readmissions can result from surgical complications, inadequate post-operative care, or socioeconomic barriers that affect follow-up adherence and recovery. Understanding factors influencing these healthcare quality measures is essential for optimizing surgical outcomes and reducing disparities in post-bariatric surgery care.

Length of hospital stay and readmission rates are particularly important metrics in bariatric surgery due to their implications for patient safety, recovery, and healthcare resource utilization(8–10). Prolonged hospital stay following bariatric surgery is often indicative of postoperative complications such as bleeding, infection, or delayed gastrointestinal function, and is associated with increased healthcare costs and poorer patient satisfaction(8, 11). However, some extended stays may be preventable with enhanced recovery protocols, multidisciplinary care teams, and improved perioperative monitoring(12). Similarly, readmissions reported in 5– 15% of bariatric cases can result from surgical site infections, anastomotic leaks, dehydration, nutritional deficiencies, and poorly managed comorbidities(8, 13, 14). Many of these readmissions may be mitigated through better discharge planning, early post-discharge follow-up, patient education, and robust outpatient support systems(14).

Beyond individual-level factors, neighborhood socioeconomic status (nSES) is an independent determinant of health outcomes(15, 16). Neighborhood socioeconomic characteristics may include income distribution, employment rates, housing stability, and healthcare accessibility(17, 18) Studies have shown that patients from low-nSES neighborhoods have longer hospital stays and higher readmission rates following various surgical procedures, including cardiac, orthopedic, and oncologic surgeries(19–21). These disparities may be driven by higher baseline comorbidities, limited healthcare resources, and increased psychosocial stressors in socioeconomically disadvantaged neighborhoods(22, 23). Despite growing recognition of the impact of nSES on surgical outcomes, research on its influence in bariatric surgery patients within state-specific healthcare models remains limited.

Several composite indices have been developed to quantify neighborhood disadvantage, each incorporating different socioeconomic factors. The Area Deprivation Index (ADI) is a census tract-based measure(24) that ranks neighborhood deprivation by considering education, income, employment, and housing quality(25). The Social Vulnerability Index (SVI), developed by the CDC, evaluates community vulnerability to external stressors by incorporating socioeconomic status, household composition, minority status, and housing characteristics(26). In contrast, the Distressed Communities Index (DCI) is a ZIP-code-based measure developed by the Economic Innovation Group, which provides a broader economic assessment of neighborhood distress by incorporating factors such as unemployment rates, poverty levels, business decline, and housing conditions(27, 28). While each of these indices offers valuable insights, the DCI captures a more comprehensive picture of economic resilience and structural barriers within communities, making it particularly useful for evaluating healthcare disparities and outcomes(24, 29). The DCI is particularly advantageous for assessing the impact of nSES on healthcare outcomes, providing a more comprehensive and dynamic reflection of economic advantages compared to ADI and SVI(24, 27).

The intersection of race/ethnicity and neighborhood disadvantage may further exacerbate disparities in post-operative outcomes(30, 31). Minority patients, particularly Black and Hispanic individuals living in low-nSES neighborhoods, might be at increased risk for prolonged hospital stays and higher readmissions due to the compounded effects of systemic barriers, lower healthcare access, and implicit biases in clinical decision-making(32–34). This intersectionality highlights the need to examine racial/ethnic disparities within the broader context of neighborhood socioeconomic disadvantages to fully understand disparities in bariatric surgery outcomes.

This study aims to evaluate the independent association between nSES (measured using the DCI) and healthcare quality measures (hospital length of stay and readmissions) following bariatric surgery in Maryland. In addition, it seeks to assess the moderating role of race/ethnicity in the relationship between nSES and post-operative outcomes. Given Maryland’s unique All-Payer Model, which standardizes hospital payment rates across all payers, this study provides a distinct opportunity to investigate healthcare disparities in a regulated reimbursement environment.

## METHODOLOGY

### Study Design and Population

This study employs a retrospective cohort design using data from the Maryland State Inpatient Database (SID) from 2018 to 2020(35). The study population includes all adult patients (≥18 years) who underwent bariatric surgery, identified using ICD-10 procedure codes for sleeve gastrectomy, gastric bypass, and other bariatric procedures. Maryland SID provides a comprehensive, statewide dataset capturing hospital discharges across all payers under the Maryland All-Payer Model, ensuring reduced bias related to insurance status.

### Primary Outcomes

The primary outcomes for this study include hospital length of stay (LOS) and readmission rates. Hospital LOS is measured as a continuous variable, representing the number of inpatient days following bariatric surgery. The second outcome, readmission, is defined as a binary variable (Yes/No), indicating whether a patient was readmitted within 30 days of discharge.

### Main Explanatory Variable: Distressed Communities Index (DCI)

Neighborhood socioeconomic status (nSES) was assessed using the DCI, a ZIP-code-based measure developed by the Economic Innovation Group. The DCI integrates various economic indicators, including unemployment rates, poverty levels, housing vacancy rates, and business decline, to provide a comprehensive assessment of economic distress at the community level. For this study, DCI was categorized into quintiles, allowing for a gradient analysis of economic disadvantages. The first quintile represents the least distressed communities, while the fifth quintile represents the most distressed neighborhoods.

### Covariates

Several demographics, clinical, socioeconomic, and geographic covariates were included in the analysis to account for potential confounders. Demographic variables included age, sex, and race/ethnicity, with racial/ethnic groups categorized as White, Black, Hispanic, and Other.

Clinical factors such as hypertension, diabetes were measured using the Charlson commodity index(36, 37), and obesity class (based on BMI categories) were incorporated to account for underlying comorbidities that may influence post-operative outcomes. Socioeconomic factors included primary insurance type, categorized as Medicare, Medicaid, Private, or Uninsured while Geographic context was stratified as Rural, Small Town, Suburban and Urban.

### Statistical Analysis

All statistical analyses were conducted using Stata 17 utilizing complete case analysis to account for missing data. Descriptive statistics were used to summarize patient characteristics, with means and standard deviations (SD) reported for continuous variables and proportions calculated for categorical variables. Chi-square tests and t-tests were used to compare baseline characteristics across DCI quintiles.

To assess the association between nSES and healthcare quality metrics, multivariable regression models were employed. For hospital length of stay, a linear regression model was used, adjusting for demographic, clinical, socioeconomic, and geographic covariates. The model was specified as follows:

LOSi=β0+β1(DCI)+β2(Covariates)

For readmission rates, a logistic regression model was applied, with results reported as adjusted odds ratios (OR) with 95% confidence intervals (CI):

Logit (P(Y=1)) =α+β 1(DCI) +β 2(Covariates)

Where: OLS for hospital stay (continuous) and a binary logistic regression for readmissions indicating whether a patient experienced a readmission following surgery (1 = Yes, 0 = No).

DCI: Main explanatory variable; α: Intercept term.;

Interaction terms were included to evaluate whether the relationship between nSES and postsurgical outcomes was modified by race/ethnicity:

Logit(P(Y=1))=α+β1(DCI)+β2(Covariates)+β3(Race)+β13(Race X DCI)

Interaction Term (β13 (Race X DCI): This term captures how the effect of the DCI on the likelihood of the outcomes (hospital stay or readmission) following surgery changes depending on the individual’s race/ethnicity. If significant, it indicates that the impact of DCI on these outcomes varies by racial group.

### Sensitivity Analysis Using Alternative Socioeconomic Index

To validate the robustness of the findings, a sensitivity analysis was performed using the Area Deprivation Index (ADI), an alternative socioeconomic measure based on census tract data. Unlike the ZIP-code-based DCI, the ADI incorporates education, income, employment, and housing characteristics to assess neighborhood deprivation. By comparing models that used DCI versus ADI, this study evaluated whether ZIP-code-based socioeconomic distress (DCI) provided a more meaningful distinction in predicting bariatric surgery outcomes compared to a censustract-based approach. This sensitivity analysis strengthened the study’s conclusions regarding the impact of neighborhood disadvantage on healthcare quality metrics.

### Ethical Considerations

This study utilized de-identified secondary data from the Maryland SID, which is exempt from direct patient consent under HIPAA regulations. The research protocol was reviewed and approved by the University of Maryland Institutional Review Board (2284677–1).

## RESULTS

The study included 10,784 adults who underwent bariatric surgery in Maryland between 2018 and 2020, distributed across five DCI quintiles (Table 1). The mean age of the cohort was 44.1 ± 11.6 years, with significant variation by DCI Quintiles. Patients from prosperous neighborhoods were older on average (mean: 45.2 years), while those from distressed areas were younger (mean: 42.7 years). The proportion of younger adults (18–44 years) was highest in distressed neighborhoods (57.3%) and lowest in prosperous areas (48.0%). Conversely, older adults (≥65 years) were more prevalent in prosperous neighborhoods (5.5%) than in distressed ones (2.8%).

**Table 1:** Baseline Characteristics of Patients Undergoing Bariatric Surgery by Distressed Communities Index Quintiles, Maryland 2018–2020.

|  | Total Population<br>(N=10,784) | Prosperous<br>(n=2,645) | Comfortable<br>(n=3,085) | Mid-Tier<br>(n=2,204) | At-Risk<br>(n=1,487) | Distressed<br>(n=1,363) | pvalue |
| --- | --- | --- | --- | --- | --- | --- | --- |
| **Age (mean ±std)** | 44.1 ±11.6 | 45.2 ±11.8 | 44.6 ±11.6 | 43.3 ±11.3 | 43.4 ±11.9 | 42.7 ±11.4 | <0.001 |
| **Age (Yr.)** |  |  |  |  |  |  | <0.001 |
| 18-44Yr. | 5,746 (52.2%) | 1,317 (48.0%) | 1,568 (49.8%) | 1219 (54.4%) | 857 (56.8%) | 785 (57.3%) |  |
| 45/64Yr. | 4,766 (43.3%) | 1,273 (46.4%) | 1,415 (45.0%) | 947 (42.2%) | 583 (38.6%) | 548 (40.0%) |  |
| >64Yr. | 501 (4.6%) | 152 (5.5%) | 165 (5.2%) | 77 (3.4%) | 69 (4.6%) | 38 (2.8%) |  |
| **Female (ref. Man)** | 9,147 (83.1%) | 2,140 (78.1%) | 2,605 (82.8%) | 1912 (85.2%) | 1309 (86.8%) | 1181 (86.1%) | <0.001 |
| **Race/Ethnicity** |  |  |  |  |  |  | <0.001 |
| White | 4,834 (44.8%) | 1,638 (61.9%) | 1,478 (47.9%) | 767 (34.8%) | 610 (41.0%) | 11 (0.8%) |  |
| Black | 5,209 (48.3%) | 812 (30.7%) | 1,391 (45.1%) | 1,243 | 779 (52.4%) |  |  |
| Hispanic | 503 (4.7%) | 111 (4.2%) | 137 (4.4%) | 157 (7.1%) | 71 (4.8%) |  |  |
| Other | 238 (2.2%) | 84 (3.2%) | 79 (2.6%) | 37 (1.7%) | 27 (1.8%) |  |  |
| **Insurance** |  |  |  |  |  |  | <0.001 |
| Self-Pay | 33 (0.3%) | 9 (0.3%) | 13 (0.4%) | 3 (0.1%) | 6 (0.4%) | 2 (0.2%) |  |
| Medicare | 988 (9.0%) | 217 (7.9%) | 289 (9.2%) | 189 (8.4%) | 134 (8.9%) | 159 (11.6%) |  |
| Medicaid | 2,110 (19.2%) | 309 (11.3%) | 481 (15.3%) | 468 (20.9%) | 399 (26.5%) | 453 (33.0%) |  |
| Private | 7,737 (70.3%) | 2,153 (78.6%) | 2,324 (73.8%) | 1543 (68.8%) | 963 (63.9%) | 754 (55.0%) |  |
| Other | 143 (1.3%) | 53 (1.9%) | 41 (1.3%) | 40 (1.8%) | 6 (0.4%) | 3 (0.2%) |  |
| **Comorbidity Index** |  |  |  |  |  |  |  |
| CCI =0 | 3,297 (29.9%) | 769 (28.1%) | 958 (30.4%) | 704 (31.4%) | 483 (32.0%) | 383 (27.9%) |  |
| CCI= 1-2 | 3,704 (33.6%) | 916 (33.4%) | 1,055 (33.5%) | 781 (34.8%) | 499 (33.1%) | 453 (33.0%) |  |
| CCI >2 | 4,012 (36.4%) | 1,057 (38.6%) | 1,135 (36.1%) | 758 (33.8%) | 527 (34.9%) | 535 (39.0%) |  |
| **Obesity Classification** |  |  |  |  |  |  |  |
| Class I Obesity | 2,401 (21.8%) | 678 (24.7%) | 754 (24.0%) | 443 (19.8%) | 292 (19.4%) | 234 (17.1%) |  |
| Class II Obesity | 8,612 (78.2%) | 2,064 (75.3%) | 2,394 (76.1%) | 1,800 (80.3%) | 1217 (80.7%) | 1137 (82.9%) | 0.006 |
| **Urban** |  |  |  |  |  |  |  |
| Rural | 1,016 (9.2%) | 357 (13.0%) | 261 (8.3%) | 197 (8.8%) | 169 | 32 (2.3%) |  |
| Small Town | 775 (7.0%) | 176 (6.4%) | 270 (8.6%) | 100 (4.5%) | 22 (1.5%) | 207 (15.1%) |  |
| Suburban | 7,426 (67.4%) | 2,051 (74.8%) | 2,518 (80.0%) | 1,668 | 1,055 | 134 (9.8%) | <0.001 |
| Urban | 1,796 (16.3%) | 158 (5.8%) | 99 (3.1%) | 278 (12.4%) | 263 | 998 (72.8%) |  |

*Baseline demographic, clinical, and socioeconomic characteristics of patients undergoing bariatric surgery in Maryland, 2018–2020, stratified by Distressed Communities Index (DCI) quintiles. Values are presented as counts (n) with percentages (%) or means with standard deviations (SD), as appropriate. Comparisons across DCI groups were assessed using chi-square tests for categorical variables and analysis of variance (ANOVA) for continuous variables.*

Women constituted 83.1% of the total population, with the highest proportions observed in atrisk (86.8%) and distressed (86.1%) communities. There were significant racial and ethnic differences by DCI quintile: White individuals represented the majority in prosperous neighborhoods (61.9%), while Black individuals were most prevalent in distressed areas (52.4%). Hispanic and other racial groups were more evenly distributed across the quintiles, although Hispanic patients had slightly higher representation in mid-tier and at-risk neighborhoods (Table 1).

Insurance coverage also demonstrated a clear socioeconomic gradient. Private insurance was more common in prosperous neighborhoods (78.6%) and decreased across quintiles, reaching 55.0% in distressed communities. Medicaid coverage followed the opposite trend, increasing from 11.3% in prosperous areas to 33.0% in distressed neighborhoods. Medicare coverage and self-pay rates remained relatively stable across groups.

The Charlson Comorbidity Index (CCI) showed that patients from distressed areas had the highest proportion with a CCI >2 (39.0%), indicating greater comorbidity burden. Obesity class also varied across quintiles: Class III obesity was more common in lower SES areas, increasing from 75.3% in prosperous to 82.9% in distressed neighborhoods (Table 1).

Geographic distribution differed significantly by SES. Patients from distressed communities were overwhelmingly urban (72.8%), while those from prosperous neighborhoods were more likely suburban (74.8%) or rural (13.0%). These findings highlight substantial demographic, clinical, and geographic disparities in bariatric surgery recipients across socioeconomic gradients, emphasizing the need for tailored public health and clinical interventions to address the distinct needs of disadvantaged populations (Table 1).

After adjusting for age, sex, insurance type, comorbidities, obesity classification, and urban categories, neighborhood socioeconomic status as measured by the DCI was not significantly associated with hospital length of stay following bariatric surgery (Table 2). Compared to individuals from Prosperous communities, those from Comfortable (β = 0.011; 95% CI: –0.091 to 0.113; p = 0.833), Mid-Tier (β = 0.055; 95% CI: –0.057 to 0.167; p = 0.334), At-Risk (β = – 0.017; 95% CI: –0.142 to 0.109; p = 0.792), and Distressed (β = –0.048; 95% CI: –0.205 to 0.108; p = 0.547) communities had no statistically significant differences in length of hospital stay (Table 2).

**Table 2:** Adjusted Association between Community Distress, Race, and Length of Stay.

| Length of Hospital Stay | Coef. | 95% CI |  | P>t |
| --- | --- | --- | --- | --- |
| **Distressed Communities Index** |  |  |  |  |
| Prosperous | Reference |  |  |  |
| Comfortable | 0.011 | -0.091 | 0.113 | 0.833 |
| Mid-Tier | 0.055 | -0.057 | 0.167 | 0.334 |
| At-Risk | -0.017 | -0.142 | 0.109 | 0.792 |
| Distressed | -0.048 | -0.205 | 0.108 | 0.547 |
| **Race & Ethnicity** |  |  |  |  |
| White | Reference |  |  |  |
| Black | 0.085 | 0.001 | 0.169 | 0.046 |
| Hispanic | 0.036 | -0.145 | 0.218 | 0.695 |
| Other | 0.119 | -0.134 | 0.372 | 0.357 |

*Multivariable linear regression models adjusted for age, sex, insurance type, comorbidities, obesity classification, and urban category. Estimates are presented as regression coefficients ( $\beta$ ) with corresponding 95% confidence intervals (CI) and p-values. White race and Prosperous communities served as reference groups.*

Among racial and ethnic groups, Black patients had a significantly longer hospital stay compared to White patients (β = 0.085; 95% CI: 0.001 to 0.169; p = 0.046). No significant differences in length of stay were observed for Hispanic (β = 0.036; 95% CI: –0.145 to 0.218; p = 0.695) or Other race patients (β = 0.119; 95% CI: –0.134 to 0.372; p = 0.357). These findings suggest that, while

Marginal effects analysis was conducted to assess whether race moderated hospital length of stay following bariatric surgery across different levels of neighborhood socioeconomic distress (Table 4)

**Table 3:** Adjusted Association between Community Distress, Race, and Readmission.

| Readmission | Odds Ratio | 95% CI |  | P>z |
| --- | --- | --- | --- | --- |
| **Distressed Communities Index** |  |  |  |  |
| Prosperous | Reference |  |  |  |
| Comfortable | 1.52 | 0.841 | 2.748 | 0.165 |
| Mid-Tier | 1.739 | 0.928 | 3.258 | 0.084 |
| At-Risk | 1.209 | 0.581 | 2.518 | 0.611 |
| Distressed | 1.409 | 0.649 | 3.059 | 0.386 |
| **Race & Ethnicity** |  |  |  |  |
| White | Reference |  |  |  |
| Black | 0.767 | 0.489 | 1.204 | 0.249 |
| Hispanic | 1.39 | 0.576 | 3.353 | 0.464 |
| Other | 0.439 | 0.06 | 3.228 | 0.418 |

*Multivariable linear regression models adjusted for age, sex, insurance type, comorbidities, obesity classification, and urban category. Estimates are presented as regression coefficients ( $\beta$ ) with corresponding 95% confidence intervals (CI) and p-values. White race and Prosperous communities served as reference groups.*

**Table 4:**
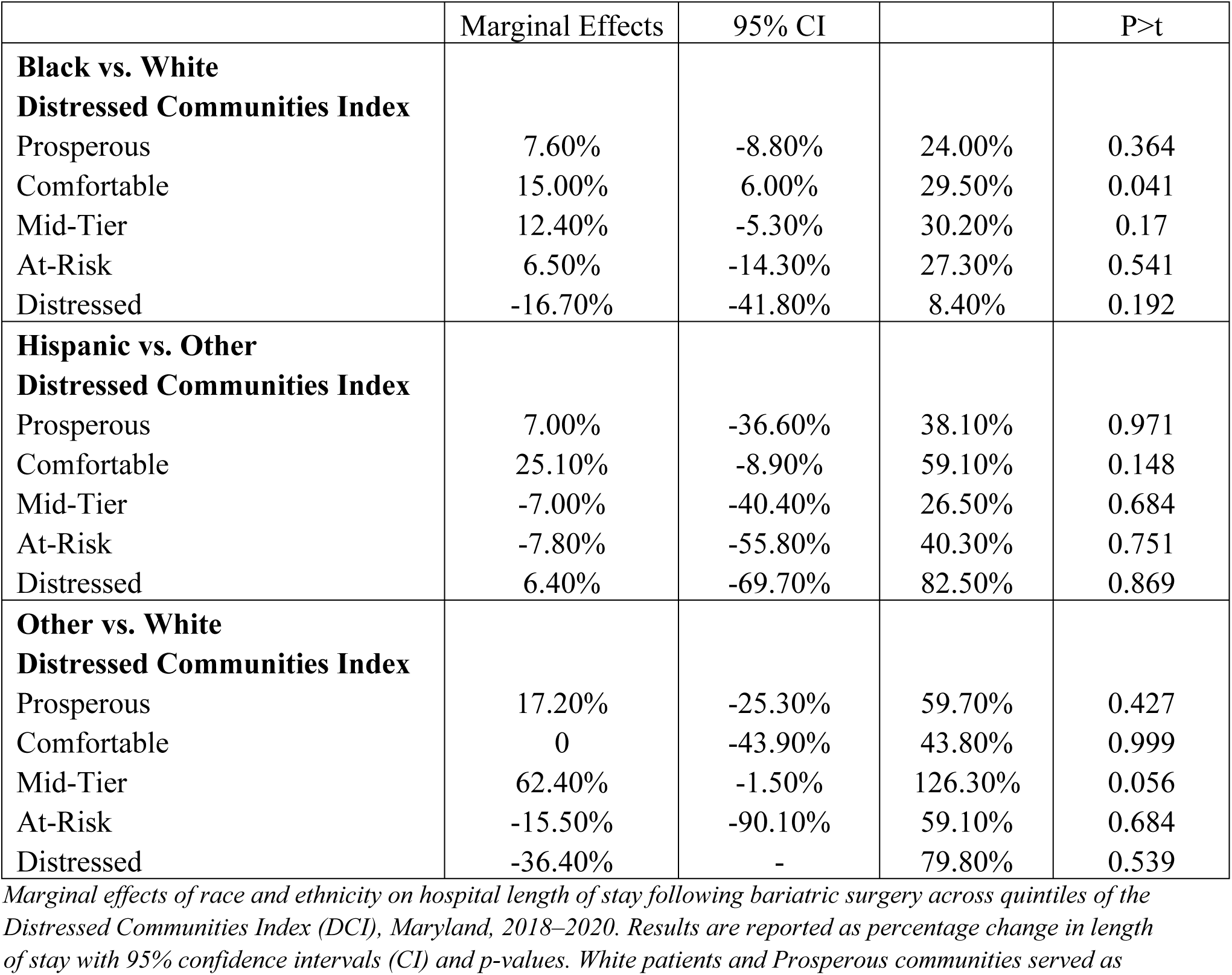
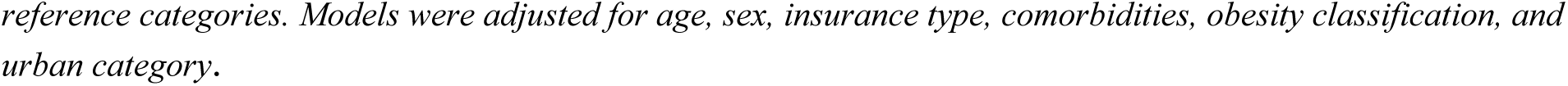
Marginal Effects of Race on Hospital Stay following Bariatric Surgery by DCI, Maryland 2018– 2020.

|  | Marginal Effects | 95% CI |  | P>t |
| --- | --- | --- | --- | --- |
| **Black vs. White** |  |  |  |  |
| **Distressed Communities Index** |  |  |  |  |
| Prosperous | 7.60% | -8.80% | 24.00% | 0.364 |
| Comfortable | 15.00% | 6.00% | 29.50% | 0.041 |
| Mid-Tier | 12.40% | -5.30% | 30.20% | 0.17 |
| At-Risk | 6.50% | -14.30% | 27.30% | 0.541 |
| Distressed | -16.70% | -41.80% | 8.40% | 0.192 |
| **Hispanic vs. Other** |  |  |  |  |
| **Distressed Communities Index** |  |  |  |  |
| Prosperous | 7.00% | -36.60% | 38.10% | 0.971 |
| Comfortable | 25.10% | -8.90% | 59.10% | 0.148 |
| Mid-Tier | -7.00% | -40.40% | 26.50% | 0.684 |
| At-Risk | -7.80% | -55.80% | 40.30% | 0.751 |
| Distressed | 6.40% | -69.70% | 82.50% | 0.869 |
| **Other vs. White** |  |  |  |  |
| **Distressed Communities Index** |  |  |  |  |
| Prosperous | 17.20% | -25.30% | 59.70% | 0.427 |
| Comfortable | 0 | -43.90% | 43.80% | 0.999 |
| Mid-Tier | 62.40% | -1.50% | 126.30% | 0.056 |
| At-Risk | -15.50% | -90.10% | 59.10% | 0.684 |
| Distressed | -36.40% | - | 79.80% | 0.539 |

*Marginal effects of race and ethnicity on hospital length of stay following bariatric surgery across quintiles of the Distressed Communities Index (DCI), Maryland, 2018–2020. Results are reported as percentage change in length of stay with 95% confidence intervals (CI) and p-values. White patients and Prosperous communities served as*
*reference categories. Models were adjusted for age, sex, insurance type, comorbidities, obesity classification, and urban category.*

Compared to White patients, Black patients had significantly longer hospital stays in Comfortable communities (marginal effect: +15.0%, 95% CI: 6.0% to 29.5%, p = 0.041). In other DCI quintiles, the differences in length of stay for Black patients were not statistically significant, with marginal effects ranging from –16.7% in Distressed areas to +12.4% in Mid-Tier communities, all with wide confidence intervals that crossed zero (Table 4).

Among Hispanic patients, none of the comparisons across DCI quintiles reached statistical significance. The marginal effects ranged from –7.8% in At-Risk communities to +25.1% in Comfortable neighborhoods (95% CI: –8.9% to 59.1%, p = 0.148), indicating considerable variability but no consistent pattern of difference in hospital stay relative to White patients (Table 4).

For patients categorized as other race, a borderline significant increase in hospital stay was observed in Mid-Tier communities (marginal effect: +62.4%, 95% CI: –1.5% to 126.3%, p = 0.056). In all other DCI quintiles, marginal effects for other race were non-significant, with wide confidence intervals, including negative values, highlighting substantial uncertainty in the estimates (Table 4).

The marginal effects analysis evaluated whether race (Black vs. White) moderated the likelihood of hospital readmission following bariatric surgery across DCI quintiles (Table 5). Overall, the differences in readmission rates between Black and White patients were small and not statistically significant across all community distress levels. In Prosperous communities, the marginal difference was +0.3% (95% CI: –0.5% to 1.1%, p = 0.51), while in Comfortable communities, the estimate was –0.5% (95% CI: –1.3% to 0.4%, p = 0.26).

**Table 5:** Marginal Effects of Race (Black vs. White) on Hospital Stay following Bariatric Surgery by DCI, Maryland 2018–2020.

| Distressed Communities Index | Marginal Effects | 95% CI |  | p-value |
| --- | --- | --- | --- | --- |
| Prosperous | 0.30% | -0.50% | 1.10% | 0.51 |
| Black vs. White |  |  |  |  |
| Comfortable | -0.50% | -1.30% | -0.40% | 0.26 |
| Mid-Tier | -0.90% | -2.00% | 0.30% | 0.13 |
| At-Risk | 0.00% | -0.90% | 1.00% | 0.92 |
| Distressed | -0.40% | -1.50% | 0.80% | 0.49 |

*Marginal effects of race (Black vs. White) on hospital readmissions following bariatric surgery across quintiles of the Distressed Communities Index (DCI), Maryland, 2018–2020. Results are reported as percentage point differences in readmission probability with 95% confidence intervals (CI) and p-values. White patients and Prosperous communities served as reference categories. Models were adjusted for age, sex, insurance type, comorbidities, obesity classification, and urban category. Due to limited sample size, marginal effects could not be calculated for other racial/ethnic groups.*

Patients from Mid-Tier and Distressed communities showed slightly lower readmission probabilities for Black patients compared to White patients (marginal effects: –0.9% and –0.4%, respectively), but confidence intervals again crossed zero, indicating non-significance. The At Risk quintile showed no difference (0.0%; 95% CI: –0.9% to 1.0%, p = 0.92). These findings suggest that race did not significantly influence readmission risk following bariatric surgery, regardless of neighborhood socioeconomic status (Table 5).

## DISCUSSION

This study examined the relationship between neighborhood socioeconomic status (nSES) and key bariatric surgery outcomes, including readmission rates and hospital length of stay. Our initial hypothesis was that lower nSES would be associated with higher readmission rates and longer hospital stays due to systemic barriers affecting post-surgical recovery, such as reduced access to healthcare resources and support services. There was no statistically significant association between neighborhood socioeconomic status, as measured by the DCI, and either readmission rates or length of hospital stay following bariatric surgery after adjusting for relevant covariates. Sensitivity analyses using the ADI reinforced these findings, suggesting that neighborhood disadvantage alone may not be a primary driver of post-surgical outcomes. Instead, factors such as hospital quality, access to post-operative care, and systemic healthcare policies may mitigate the expected disparities(38–40).

The lack of a statistically significant association may be influenced by Maryland’s All-Payer Model, which standardizes hospital reimbursement rates, potentially reducing financial barriers to care(41, 42). In addition, the presence of bariatric centers of excellence in the state may contribute to uniform surgical protocols and post-operative management, ensuring more equitable outcomes across socioeconomic groups(25, 43, 44). While lower socioeconomic status has been linked to worse surgical outcomes in prior research(45, 46), some studies consistent with ours have found that these associations weaken when hospital-level factors and policy-driven interventions are considered(45, 47–49). Future research should explore the role of individual-level socioeconomic factors, healthcare literacy, and long-term adherence to post-surgical care in shaping bariatric surgery outcomes.

Race and ethnicity did not significantly modify the association between nSES and surgical outcomes, suggesting that broader structural determinants of health, such as access to care, provider networks, and community resources, may play a more influential role in post-surgical recovery than individual racial identity(50, 51). Standardized bariatric surgery protocols and postoperative care guidelines may also help mitigate disparities by ensuring consistent preoperative education, surgical management, and follow-up care(52, 53). However, unmeasured factors, such as implicit biases in healthcare delivery and differences in post-surgical adherence, may still contribute to disparities(54). Addressing systemic barriers to care, including neighborhood healthcare infrastructure, insurance accessibility, and provider availability, may be more effective in reducing disparities than focusing solely on race or ethnicity.

Although our findings did not show significant associations, they underscore the complex interplay between socioeconomic status, healthcare policies, and surgical outcomes. Future research should investigate long-term weight loss maintenance, complication rates, and healthcare utilization patterns to better understand how social disadvantage manifests in bariatric surgery outcomes.

### Policy and Research Implications

From a policy perspective, our findings suggest that neighborhood disadvantage alone may not be a sufficient predictor of post-surgical outcomes, particularly in healthcare systems implementing strong policy interventions such as Maryland’s All-Payer Model. Nonetheless, the observed trends of higher readmission rates and prolonged hospital stays among patients from distressed communities underscore the ongoing influence of socioeconomic disparities though these may operate through more nuanced mechanisms. As the landscape of obesity treatment evolves with the growing availability of GLP-1 receptor agonists, such as semaglutide and tirzepatide, it is essential to contextualize the role of bariatric surgery within a broader treatment framework. While GLP-1s have demonstrated significant effectiveness in promoting weight loss and improving metabolic parameters, access remains limited by cost, insurance coverage, and long-term adherence challenges, particularly in underserved populations. Therefore, bariatric surgery should continue to be promoted as a viable and often necessary option for managing class II and III obesity, especially for patients who may not respond adequately to or cannot access pharmacologic therapies.

Future research should focus on integrating individual-level socioeconomic data with neighborhood-level indicators to better understand the pathways through which disadvantage affects surgical outcomes. Furthermore, targeted interventions to enhance post-surgical care adherence, improve patient education, and reduce barriers to follow-up especially in underserved communities are warranted. Nationally representative, longitudinal studies are needed to assess how emerging pharmacologic options and persistent socioeconomic disparities intersect to influence long-term outcomes in the management of severe obesity.

### Study Limitations

This study has several limitations that should be acknowledged. First, while we utilized robust socioeconomic measures such as the DCI and ADI, these are neighborhood-level indicators and may not fully capture individual-level socioeconomic factors such as income, education, or healthcare access, which could independently influence hospital readmissions and length of stay. Second, our analysis was conducted within Maryland’s All-Payer Model, a unique healthcare system that may reduce socioeconomic disparities in access to care, potentially limiting the generalizability of our findings to other states with different healthcare reimbursement structures.

In addition, while our study suggests a trend toward higher readmissions and prolonged hospital stays in distressed communities, the lack of statistical significance may reflect insufficient power, residual confounding, or unmeasured variables, such as post-discharge care access and patient adherence to follow-up recommendations. Lastly, our reliance on administrative claims data introduces the potential for misclassification bias, as certain clinical and social factors influencing surgical outcomes (e.g., nutrition status, social support, and health literacy) are not captured in hospital discharge records. Future research should incorporate individual-level socioeconomic data, patient-reported outcomes, and qualitative assessments to better elucidate the mechanisms driving disparities in post-bariatric surgery outcomes.

### Conclusion

In conclusion, our study found no statistically significant association between neighborhood socioeconomic status, as measured by the DCI, and either readmission rates or length of hospital stay following bariatric surgery. These findings suggest that neighborhood-level disadvantage, as captured by the DCI, may not independently predict these specific healthcare quality outcomes in a regulated healthcare environment such as Maryland’s All-Payer Model. Nonetheless, neighborhood socioeconomic status remains a relevant contextual factor for understanding broader disparities in surgical care. Continued research is warranted to explore other structural and individual-level determinants that may contribute to variations in post-operative recovery and to inform targeted interventions for at-risk populations.

## Data Availability

The data underlying the results of this study are available from the corresponding author upon reasonable request.

## References

1. Gulinac M, Miteva DG, Peshevska-Sekulovska M, Novakov IP, Antovic S, Peruhova M, et al. Long-term effectiveness, outcomes and complications of bariatric surgery. World J Clin Cases. 2023;11(19):4504–12.

2. Akinyemi OA, Weldeslase TA, Fasokun M, Griffiths Y, Andine T, Odusanya E, et al. The impact of the affordable care act on access to bariatric surgery in Maryland. Am J Surg. 2024;235:115609.

3. Noria SF, Grantcharov T. Biological effects of bariatric surgery on obesity-related comorbidities. Can J Surg. 2013;56(1):47–57.

4. Stefan MS, Pekow PS, Nsa W, Priya A, Miller LE, Bratzler DW, et al. Hospital performance measures and 30-day readmission rates. J Gen Intern Med. 2013;28(3):377–85.

5. Chopra I, Wilkins TL, Sambamoorthi U. Hospital length of stay and all-cause 30-day readmissions among high-risk Medicaid beneficiaries. J Hosp Med. 2016;11(4):283–8.

6. Ranathunga L. Prolonged Hospital Stays, and Their Impact On Health Care Resources: Premier Health; 2024 [Available from: https://www.premierhealth.com/your-health/articles/premier-pulse/prolonged-hospital-stays--and-their-impact-on-health-care-resources.

7. Jenkins TM, Sharrack B. The cost of long hospital stays. Clin Med (Lond). 2012;12(1):98–9.

8. Lois AW, Frelich MJ, Sahr NA, Hohmann SF, Wang T, Gould JC. The relationship between duration of stay and readmissions in patients undergoing bariatric surgery. Surgery. 2015;158(2):501–7.

9. Vierra BM, Edgerton CA, Shikora SA. The impact of procedure type on 30-day readmissions following metabolic and bariatric surgery: postoperative complications of bariatric surgery. Surg Endosc. 2023;37(3):2127–32.

10. Quadri P, Sanchez-Johnsen L, Aguiluz-Cornejo G, Masrur M, Sigmon D, Danielson KK, et al. Bariatric Surgery Hospital Readmissions in an Urban Academic Medical Center. Bariatric Surgical Practice and Patient Care. 2023;18(2):80–4.

11. Nijland LMG, de Castro SMM, van Veen RN. Risk Factors Associated with Prolonged Hospital Stay and Readmission in Patients After Primary Bariatric Surgery. Obes Surg. 2020;30(6):2395–402.

12. Major P, Wysocki M, Torbicz G, Gajewska N, Dudek A, Małczak P, et al. Risk Factors for Prolonged Length of Hospital Stay and Readmissions After Laparoscopic Sleeve Gastrectomy and Laparoscopic Roux-en-Y Gastric Bypass. Obes Surg. 2018;28(2):323–32.

13. Dang JT, Tavakoli I, Switzer N, Mocanu V, Shi X, de Gara C, et al. Factors that predict 30-day readmission after bariatric surgery: experience of a publicly funded Canadian centre. Can J Surg. 2020;63(2):E174–e80.

14. Sharma P. Reducing Early Hospital Readmission Rates after Bariatric Surgery: ProQuest Dissertations & Theses; 2021.

15. DeVille NV, Iyer HS, Holland I, Bhupathiraju SN, Chai B, James P, et al. Neighborhood socioeconomic status and mortality in the nurses’ health study (NHS) and the nurses’ health study II (NHSII). Environmental Epidemiology. 2023;7(1):e235.

16. Topel ML, Kim JH, Mujahid MS, Sullivan SM, Ko YA, Vaccarino V, et al. Neighborhood Socioeconomic Status and Adverse Outcomes in Patients With Cardiovascular Disease. Am J Cardiol. 2019;123(2):284–90.

17. Arcaya MC, Ellen IG, Steil J. Neighborhoods And Health: Interventions At The Neighborhood Level Could Help Advance Health Equity. Health Affairs. 2024;43(2):156–63.

18. Arcaya MC, Tucker-Seeley RD, Kim R, Schnake-Mahl A, So M, Subramanian SV. Research on neighborhood effects on health in the United States: A systematic review of study characteristics. Soc Sci Med. 2016;168:16–29.

19. Vashist S, Dudeck BS, Sherfy B, Rosenthal GL, Chaves AH. Neighborhood socioeconomic status and length of stay after congenital heart disease surgery. Frontiers in Pediatrics. 2023;11.

20. Zumbrunn A, Bachmann N, Bayer-Oglesby L, Joerg R, Team tS. Social disparities in unplanned 30-day readmission rates after hospital discharge in patients with chronic health conditions: A retrospective cohort study using patient level hospital administrative data linked to the population census in Switzerland. medRxiv. 2022:2022.01.18.22269480.

21. El Moheb M, Kareddy A, Young S, Weber M, Noona S, Wisniewski A, et al. Assessing the impact of socioeconomic distress on hospital readmissions after cardiac surgery. JTCVS Open. 2024;21:211–23.

22. Bonner SN, Ibrahim AM, Kunnath N, Dimick JB, Nathan H. Neighborhood Deprivation, Hospital Quality, and Mortality After Cancer Surgery. Ann Surg. 2023;277(1):73–8.

23. Dawson LP, Andrew E, Nehme Z, Bloom J, Biswas S, Cox S, et al. Association of Socioeconomic Status With Outcomes and Care Quality in Patients Presenting With Undifferentiated Chest Pain in the Setting of Universal Health Care Coverage. Journal of the American Heart Association. 2022;11(7):e024923.

24. Mehaffey JH, Hawkins RB, Charles EJ, Turrentine FE, Hallowell PT, Friel C, et al. Socioeconomic “Distressed Communities Index” Improves Surgical Risk-adjustment. Ann Surg. 2020;271(3):470–4.

25. Gray M. Pratt PG-U. Bariatric Surgery Centers of Excellence®: Why they are important when selecting your surgeon and hospital. [Available from: https://www.obesityaction.org/resources/bariatric-surgery-centers-of-excellence-why-they-are-important-when-selecting-your-surgeon-and-hospital<u>/</u>.

26. Rollings KA, Noppert GA, Griggs JJ, Melendez RA, Clarke PJ. Comparison of two area-level socioeconomic deprivation indices: Implications for public health research, practice, and policy. PLoS One. 2023;18(10):e0292281.

27. Akinyemi OA, Omokhodion OV, Fasokun ME, Makanjuola OE, Aaron S, Elleissy Nasef K, et al. Exploring the Relationship Between Community-Level Economic Deprivation and HIV Infection Among Hospital Admissions in Washington, DC. Cureus. 2023;15(4):e37236.

28. Economic Innovation Group. Distressed Communities Index Maps the Country’s Economic Well-Being: Georgia Municipal Association; 2020 [Available from: https://www.gacities.com/Resources/Reference-Articles/Distressed-Communities-Index-Maps-the-Country-s-Ec.aspx?feed=93d3a9e3-258c-4a31-8540-e69641995743.

29. Charles EJ, Mehaffey JH, Hawkins RB, Fonner CE, Yarboro LT, Quader MA, et al. Socioeconomic Distressed Communities Index Predicts Risk-Adjusted Mortality After Cardiac Surgery. Ann Thorac Surg. 2019;107(6):1706–12.

30. Vohra-Gupta S, Petruzzi L, Jones C, Cubbin C. An Intersectional Approach to Understanding Barriers to Healthcare for Women. J Community Health. 2023;48(1):89–98.

31. Vela MB, Erondu AI, Smith NA, Peek ME, Woodruff JN, Chin MH. Eliminating Explicit and Implicit Biases in Health Care: Evidence and Research Needs. Annu Rev Public Health. 2022;43:477–501.

32. Yearby R, Clark B, Figueroa JF. Structural Racism In Historical And Modern US Health Care Policy. Health Affairs. 2022;41(2):187–94.

33. Stenberg E, Näslund I, Persson C, Szabo E, Sundbom M, Ottosson J, et al. The association between socioeconomic factors and weight loss 5 years after gastric bypass surgery. International Journal of Obesity. 2020;44(11):2279–90.

34. Blair IV, Steiner JF, Havranek EP. Unconscious (implicit) bias and health disparities: where do we go from here? Perm J. 2011;15(2):71–8.

35. Agency for Healthcare Research and Quality. SID Database Documentation; State Inpatient Databases (SID) Database Documentation. H-CUP; 2025.

36. Charlson ME, Carrozzino D, Guidi J, Patierno C. Charlson Comorbidity Index: A Critical Review of Clinimetric Properties. Psychotherapy and Psychosomatics. 2022;91(1):8–35.

37. Huang YQ, Gou R, Diao YS, Yin QH, Fan WX, Liang YP, et al. Charlson comorbidity index helps predict the risk of mortality for patients with type 2 diabetic nephropathy. J Zhejiang Univ Sci B. 2014;15(1):58–66.

38. Diaz A, Lindau ST, Obeng-Gyasi S, Dimick JB, Scott JW, Ibrahim AM. Association of Hospital Quality and Neighborhood Deprivation With Mortality After Inpatient Surgery Among Medicare Beneficiaries. JAMA Network Open. 2023;6(1):e2253620-e.

39. Diaz A, Beane JD, Hyer JM, Tsilimigras D, Pawlik TM. Impact of hospital quality on surgical outcomes in patients with high social vulnerability: Association of textbook outcomes and social vulnerability by hospital quality. Surgery. 2022;171(6):1612–8.

40. Marta LM, Tanya LZ, Lisa Marie K, Brandon B, Lillian SK, Kathie-Ann J, et al. Taking action to achieve health equity and eliminate healthcare disparities within acute care surgery. Trauma Surgery & Acute Care Open. 2024;9(1):e001494.

41. CMS.gov CfMMS. Maryland All-Payer Model: CMS.gov, Centers for Medicare & Medicaid Services,; [Available from: https://www.cms.gov/priorities/innovation/innovation-models/maryland-all-payer-model.

42. Douglas Holtz-Eakin AS. The National Implications of Maryland’s All-Payer System: American Action Forum; 2020 [Available from: https://www.americanactionforum.org/research/the-national-implications-of-marylands-all-payer-system/.

43. Ibrahim AM, Ghaferi AA, Thumma JR, Dimick JB. Variation in Outcomes at Bariatric Surgery Centers of Excellence. JAMA Surg. 2017;152(7):629–36.

44. Gallagher AG, Angelo RL, Kearney P. Factors Associated With Variation in Outcomes in Bariatric Surgery Centers of Excellence. JAMA. 2018;320(13):1386–7.

45. Birkmeyer NJ, Gu N, Baser O, Morris AM, Birkmeyer JD. Socioeconomic status and surgical mortality in the elderly. Med Care. 2008;46(9):893–9.

46. Williamson CG, Richardson S, Ebrahimian S, Kronen E, Verma A, Benharash P. Identifying the origin of socioeconomic disparities in outcomes of major elective operations. Surg Open Sci. 2023;13:66–70.

47. Sheetz KH, Ibrahim AM, Nathan H, Dimick JB. Variation in Surgical Outcomes Across Networks of the Highest-Rated US Hospitals. JAMA Surgery. 2019;154(6):510–5.

48. Qasim M, Andrews RM. Despite Overall Improvement In Surgical Outcomes Since 2000, Income-Related Disparities Persist. Health Affairs. 2013;32(10):1773–80.

49. Stephens TJ, Peden CJ, Haines R, Grocott MPW, Murray D, Cromwell D, et al. Hospital-level evaluation of the effect of a national quality improvement programme: time-series analysis of registry data. BMJ Quality & Safety. 2020;29(8):623–35.

50. Ostovari M, Yu D. Impact of care provider network characteristics on patient outcomes: Usage of social network analysis and a multi-scale community detection. PLoS One. 2019;14(9):e0222016.

51. Paul RW, Osman A, Nigro A, Muchintala R, Destine H, Tjoumakaris FP, et al. The effects of social determinants of health on rotator cuff repair utilization and outcomes: a systematic review. JSES Rev Rep Tech. 2024;4(3):346–52.

52. Bhandari M, Fobi MAL, Buchwald JN. Standardization of Bariatric Metabolic Procedures: World Consensus Meeting Statement. Obes Surg. 2019;29(Suppl 4):309–45.

53. Abbott S, Shuttlewood E, Flint SW, Chesworth P, Parretti HM. “Is it time to throw out the weighing scales?” Implicit weight bias among healthcare professionals working in bariatric surgery services and their attitude towards non-weight focused approaches. EClinicalMedicine. 2023;55:101770.

54. Aviva H. Ariel-Donges; Carlysa K. Oyama; and Megan M. Hood. Patient-reported Short-term Barriers to and Facilitators of Adherence to Behavioral Recommendations Following Bariatric Surgery. Bariatric Times,. 2020.

